# Performance of ChatGPT on the MCAT: The Road to Personalized and Equitable Premedical Learning

**DOI:** 10.1101/2023.03.05.23286533

**Authors:** Vikas L Bommineni, Sanaea Bhagwagar, Daniel Balcarcel, Christos Davatzikos, Donald Boyer

## Abstract

Despite an increasingly diverse population, an unmet demand for undergraduates from underrepresented racial and ethnic minority (URM) backgrounds exists in the field of medicine as a result of financial hurdles and insufficient educational support faced by URM students in the premedical journey. With the capacity to provide highly individualized and accessible no- or low-cost dynamic instruction, large language models (LLMs) and their chatbot derivatives are posed to change this dynamic and subsequently help shape a more diverse future physician workforce. While studies have established the passing performance and insightful explanations of one of the most accurate LLM-powered chatbots to date—Chat Generative Pre-trained Transformer (ChatGPT)—on standardized exams such as medical licensing exams, the role of ChatGPT in premedical education remains unknown. We evaluated the performance of ChatGPT on the Medical College Admission Test (MCAT), a standardized 230-question multiple choice exam that assesses a broad range of competencies in the natural, physical, social, and behavioral sciences as well as critical analysis and reasoning. Depending on its visual item response strategy, ChatGPT performed at or above the median performance of 276,779 student test takers on the MCAT. Additionally, ChatGPT-generated answers demonstrated both a high level of agreement with the official answer key as well as insight into its explanations. Based on these promising results, we anticipate two primary applications of ChatGPT and future LLM iterations in premedical education: firstly, such models could provide free or low-cost access to personalized and insightful explanations of MCAT competency-related questions to help students from all socioeconomic and URM backgrounds. Secondly, these models could be used to generate additional test questions by test-makers or for targeted preparation by pre-medical students. These applications of ChatGPT in premedical education could be an invaluable, innovative path forward to increase diversity and improve equity among premedical students.

## INTRODUCTION

With a rapidly diversifying population in the United States of America, there remains an unmet demand for undergraduates of underrepresented racial and ethnic minority (URM) backgrounds to pursue premedical tracks due to financial and social hurdles^1^. As a result, these URM groups continue to remain underrepresented in the physician workforce.

Several challenges to the recruitment and retention of URM undergraduate students in the premedical track contribute to the ultimate lack of diversity in matriculated medical student populations. The strong financial burden of pursuing a medical career is evident in the opportunity cost of taking rigorous science, technology, engineering, and mathematics (STEM) courses, the cost of taking the required premedical courses, and the decision to take on a six-figure debt for graduate school. These factors dissuade many students from low socioeconomic backgrounds, who disproportionately tend to be URM rather than their non-URM peers^2,3^. As importantly, there is a documented lack of advising and perceived educational support for URMs, affecting their likelihood of persistence in completing the premedical track^2,4^. Given the complexity and breadth of undergraduate coursework and examinations required to become a physician, new methods to provide personalized yet cost-effective educational assistance for these students are highly sought after. One of the most promising ways is through artificial intelligence (AI).

The application of AI in medicine, technology, and education has steadily grown in recent years. Deep learning, a subset of machine learning in which the parameters of multi-layer neural networks are trained, is responsible for many breakthroughs in processing images, speech, audio, and video^5^. The advent of deep learning algorithms referred to as large language models (LLM) has initiated questions across popular media about the ability of AI to perform critical reasoning and serve as a tool in education.

The most promising LLMs trained on corpora of texts published on the Internet can read, interpret, and generate text based on a network of interconnected tokens^6^. These tokens can be thought of as text pieces comprised of words, where user inputs are broken down into the pieces depending on how probable or frequent a token is in the corpus of training text. Due to this structure, these tools can infer relationships between words, offer predictions on tasks requiring logical language reasoning, and generate answers when prompted with personalized questions. Over the last 5 years, token-based LLMs have been released with rapidly increasing levels of capability. Three recent LLMs – Megatron Language Model (MegatronLM)^7^, Generative Pretrained Transformer 2 (GPT-2)^8^, and Turing Natural Language Generation (T-NLG)^9^ – have demonstrated exponential leaps in performance by achieving consecutively groundbreaking levels of accuracy on a variety of natural language processing tasks including question answering and language inferences. This performance gain by LLMs has stemmed from the simple scaling of data and computational resources in the form of increasing text inputs and machine learning parameters, respectively^10^. These factors have prepared LLMs to act as Internet chatbots or web-accessible computer program interfaces that simulate and process human conversations.

ChatGPT is the most recent and powerful iteration of Internet chatbots. As of early 2023, consisting of 175 billion parameters, ChatGPT is the largest trained Internet chatbot in human history, resulting in unprecedented accuracy on standardized language tasks^11^. Recent studies have applied this chatbot formulation of the finetuned GPT-3.5 model to standardized tests. While studies have established the passing performance of ChatGPT on standardized exams such as medical licensing exams^12,13^ along with business^14^ and law school^15^ exams, the role of ChatGPT in premedical education remains unknown. With its recent successful applications in professional licensing exams and its potential utility as a tool to help diversify the physician workforce, it is important to consider how LLMs, particularly ChatGPT, can affect premedical education by providing accessible low- or no-cost personalized learning.

In this study, we evaluate the performance of ChatGPT on its ability to perform critical reasoning tasks on undergraduate premedical content materials by testing its performance on questions from the Medical College Admission Test (MCAT). The MCAT is a standardized, multiple-choice examination of 230 questions taken by more than 85,000 students per year that is required for admission to medical schools in the United States and Canada. The Association of American Medical Colleges (AAMC) appoints PhD-level experts to create the exam questions to test a broad range of competencies in the natural, physical, social, and behavioral sciences as well as critical analysis and reasoning essential to entering students’ success in medical school.

The purpose of our study is to evaluate the performance of ChatGPT on the MCAT exam and determine its ability to assist students in preparation for the exam. We hypothesized that ChatGPT would outperform the median overall score and section scores of the 276,779 students who have taken the exam between the years 2019 and 2021, demonstrate high agreement with the official answer key in its explanations, and attain a high insight prevalence in its explanations. We assess the accuracy, agreement to the official AAMC answer key, and insight into ChatGPT’s responses. By providing responses that are accurate, logically concordant with the official answer key, and insightful to questions related to these courses, ChatGPT would be poised to augment students’ preparation for the MCAT exam and subsequently expand the pipeline of URM students matriculating into medical school.

## METHODS

An overview of the methodology as well as an applied example of how a passage-based question was encoded and adjudicated can be found in Figure 1.

**Figure 1.**
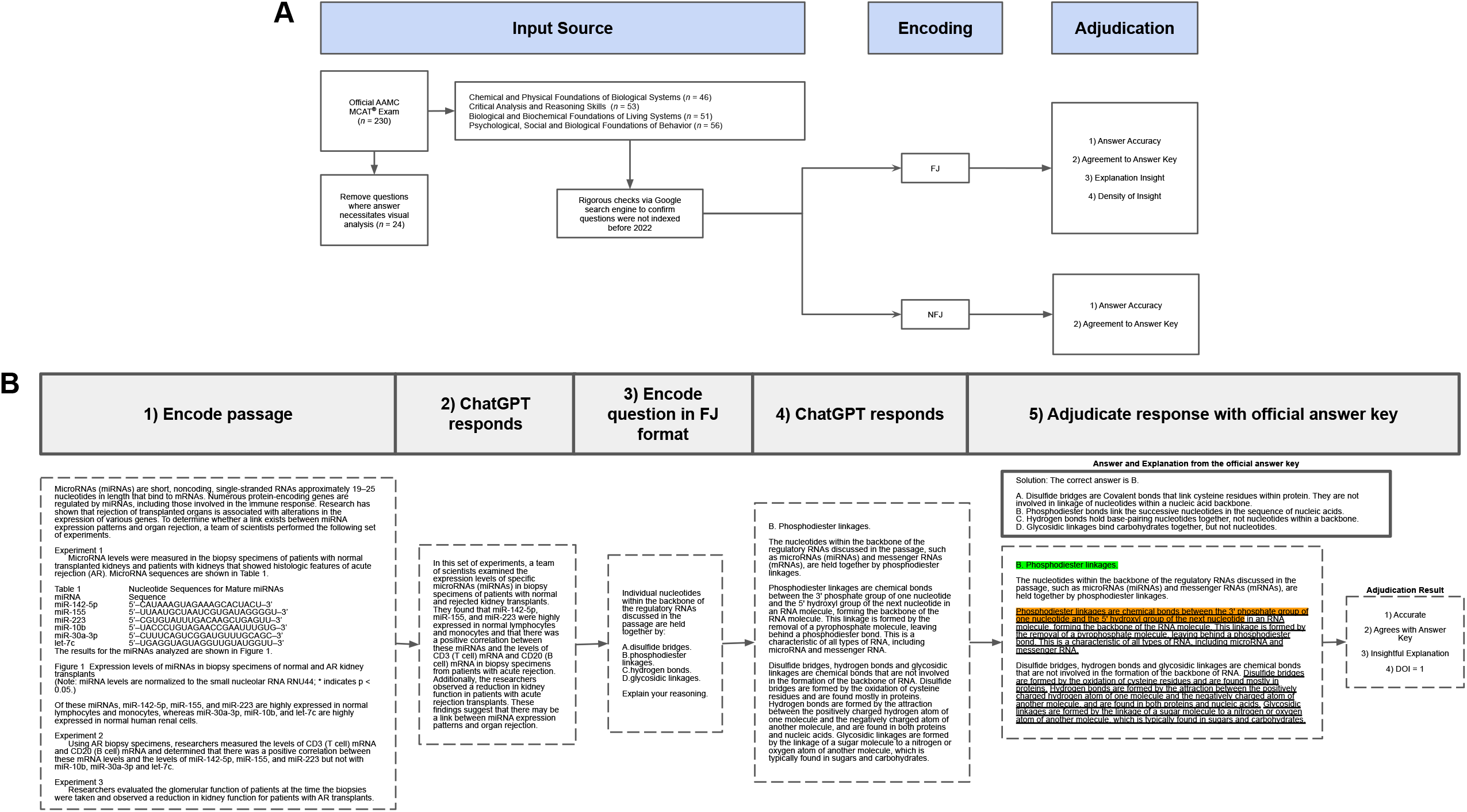
Overview of methodology workflow (A) and an example (B) of the encoding and adjudication processes of the workflow for an FJ encoded variant. The green highlighted text corresponds to the part of ChatGPT’s answer that led the adjudicator to their conclusion on “Answer Accuracy”. The orange highlighted text corresponds to the part of ChatGPT’s answer that led the adjudicator to their conclusion on “Agreement to Answer Key”. The underlined text corresponds to what part of ChatGPT’s answer led the adjudicator to their conclusions on “Explanation Insight” and “Density of Insight”. Abbreviations: FJ = multiple choice single answer with forced justification; NFJ = multiple choice single answer with no forced justification; DOI = Density of Insight; AAMC = The Association of American Medical Colleges; MCAT = Medical College Admission Test; ChatGPT = Chat Generative Pre-trained Transformer.

### Defining ChatGPT

ChatGPT is a large language model that uses advanced machine-learning techniques to understand and generate natural human language^11^. It can be used to generate text, answer questions, and carry on a conversation. Because it has been trained on a large dataset of human language, it can understand and respond to a wide range of topics and questions. Importantly, ChatGPT is a server-contained language model that is unable to inform its responses via conducting internet searches. This is different from chatbots or conversational systems that are allowed to use external sources of information, such as searching online or accessing databases, to give more informed answers to user questions.

### Input Source

230 publicly available multiple choice test questions were obtained from an October 2022 AAMC sample exam release available on the official MCAT website^16^. This freely available exam consists entirely of previously administered passage-based and free-response exam questions. Since questions from this exam had been used in previously administered exams, a rigorous check was performed to ensure that none of the answers, explanations, or related content to the questions were indexed by the Google search engine prior to January 1, 2022, representing the last date accessible to the ChatGPT training dataset. This check involved separately inputting each answer, explanation, and question into the Google search engine, followed by confirming the next 6 pages of search results did not index them before January 1, 2022. Since ChatGPT can only process text-based inquiries and encoding of visual items would introduce interpretive bias into our experiments, all sample test questions were manually screened, and questions necessitating visual analysis to answer them, such as graphs and diagrams, were removed. In all tasks except calculating a scaled score, 206 MCAT items (Chemical and Physical Foundations of Biological Systems (CP) section: 46 items, Critical Analysis and Reasoning Skills (CARS) section: 53 items, Biological and Biochemical Foundations of Living Systems (BB) section: 51 items, Psychological, Social and Biological Foundations of Behavior (PS) section: 56 items) were advanced to question encoding.

### Encoding

For passage-based questions, the passage was first inputted into ChatGPT manually, followed by the questions related to the passage; however, we removed the “Adapted From” attribution tag from the bottom of passages to prevent biasing ChatGPT to rely on out-of-scope knowledge obtained from the source text. Alternatively, for free-response questions, the questions were individually inputted into ChatGPT. A new chat session was started in ChatGPT for each passage and its associated questions as well as for each free-response question to prevent ChatGPT from using information between unconnected question groups.

To probe into ChatGPT’s ability to generate explanations, following Tseng et al’s methodology^13^, both passage-based and free-response questions were encoded into two variants and inputted into ChatGPT:

1. Multiple choice single answer without forced justification (NFJ): Created by reproducing the original MCAT question verbatim. For example: “Individual nucleotides within the backbone of the regulatory RNAs discussed in the passage are held together by: A. disulfide bridges. B. phosphodiester linkages. C. hydrogen bonds. D. glycosidic linkages.”
2. Multiple choice single answer with forced justification (FJ): Created by adding a variable lead-in or follow-on imperative or interrogative phrase asking ChatGPT to provide a rationale for each answer choice. For example: “Individual nucleotides within the backbone of the regulatory RNAs discussed in the passage are held together by: A. disulfide bridges. B. phosphodiester linkages. C. hydrogen bonds. D. glycosidic linkages. Explain your reasoning.”

A new chat was also started between the different encoding variants. Encoders employed deliberate variation in the lead-in and follow-on prompts to avoid systemic errors that could be caused by stereotyped wording as well as to simulate normal conversations.

### Adjudication

For FJ-encoded questions, ChatGPT outputs were independently judged for “Answer Accuracy”, “Agreement to the Answer Key”, “Explanation Insight”, and “Density of Insight” by an adjudicator using a rubric derived from Tseng et. al’s Accuracy-Concordance-Insight scoring system (Table 1); NFJ-encoded variants were judged for “Answer Accuracy” and “Agreement to the Answer Key” only. The “Density of Insight” metric is calculated by taking the number of ChatGPT’s explanations for each of the possible answer choices that meet the insightful criteria and dividing by 4, the number of possible answer choices on the MCAT.

**Table 1.**
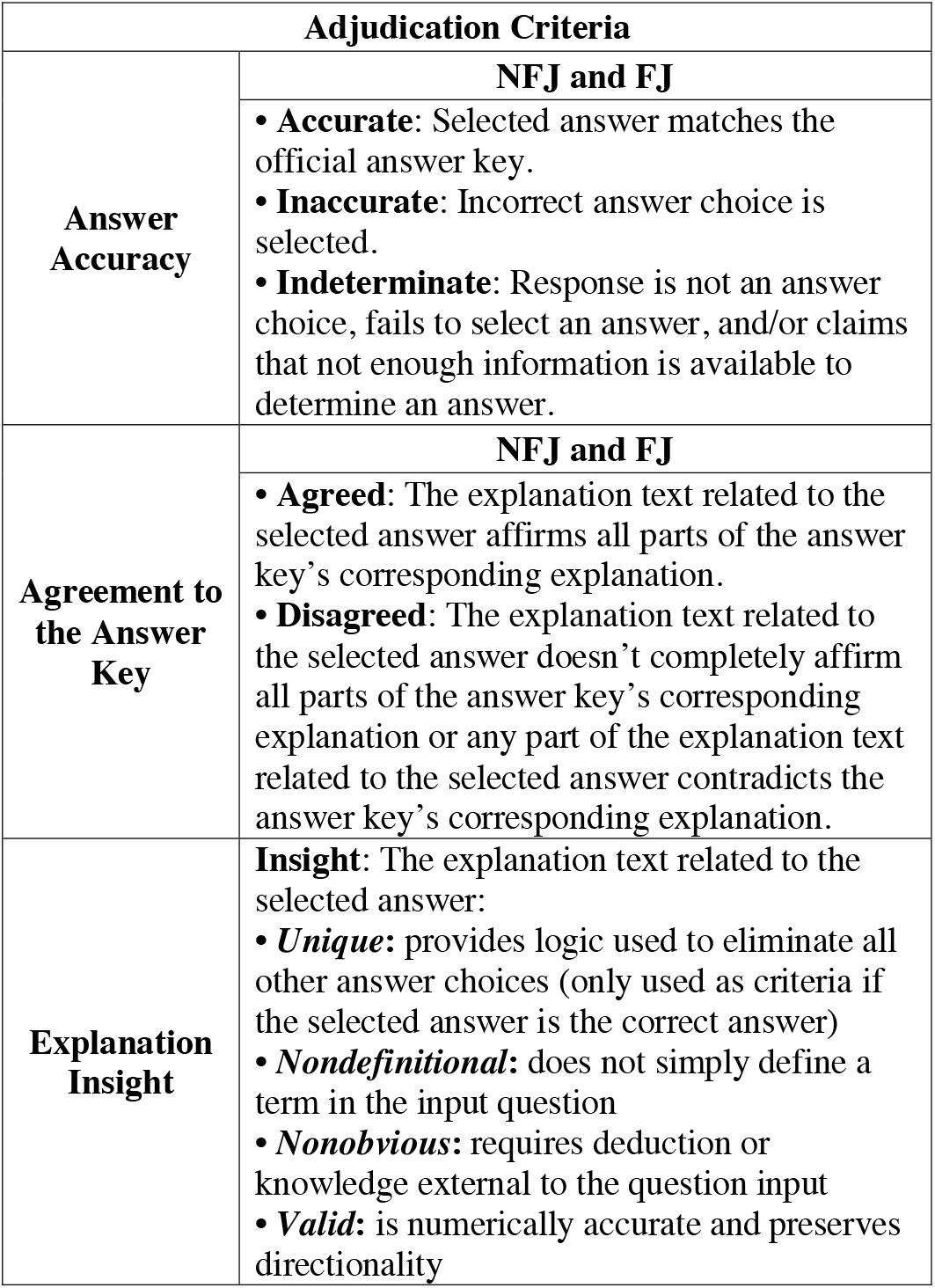
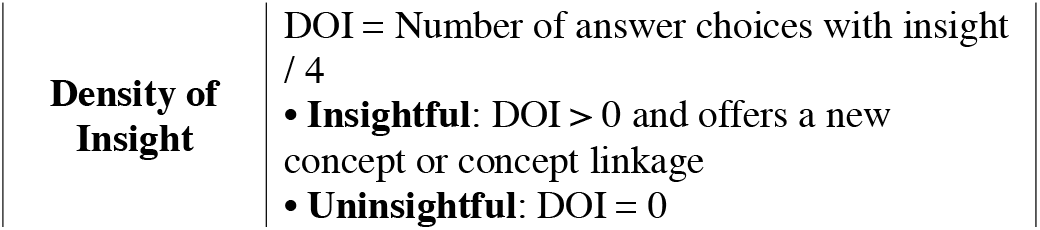
Adjudication criteria used to evaluate the answer accuracy, agreement with the answer key, insight of the explanation associated with ChatGPT’s answer, and the density of insight across all possible answers. The table is derived from Tseng et. al.^13^.

To minimize within-item anchoring bias, the adjudicator scored Answer Accuracy for all items, followed by Agreement to the Answer Key for all items, followed by Explanation Insight for all items, and lastly followed by Density of Insight.

### Scaled Scoring and Comparison Procedures

Each of the four sections on the MCAT (BB, CP, CARS, and PS) is scored from a low of 118 to a high of 132, with a theoretical median of 125. The total score is determined from the sum of the four section scores, thus ranging from a low of 472 to a high of 528 with a theoretical median of 500. Wrong answers are scored the same as unanswered questions; there is no additional penalty for wrong answers.

According to the *“How is the MCAT Exam scored?”* section on the AAMC’s website^17^, their undisclosed conversion methodology from the raw number of correct answers to these scaled scores compensates for small variations in difficulty between sets of questions. This equating process enables the generalization of our results to all MCAT exams consisting of different questions. The AAMC does not publish the raw-to-scaled scoring conversion, but it does provide individuals with a scaled score at the end of the online exam. To determine ChatGPT’s scaled performance, we inputted the raw number of questions answered correctly in each section and obtained raw-to-scaled scoring equivalences.

Then, to test our hypothesis that ChatGPT would outperform the median overall score and sections scores of students who’ve taken the exam from 2019 to 2021, we obtained the median scores (rounded to the nearest whole number to allow comparison) for all sections and the overall test was determined via percentile ranks calculated by the AAMC, based on the scores of everyone (*n* = 276,779) who tested in 2019, 2020, and 2021; however, these scores were obtained with those test-takers having the opportunity to answer the visual item questions.

To fairly evaluate the performance of ChatGPT on the entire exam and thus obtain a scaled score to compare to observed median student performance, we chose to model two scenarios on how ChatGPT could potentially respond to questions containing visual items (collectively referred to as “visual item response strategies”):

1. Answer by making an educated guess based on the article’s context (generated via prompting ChatGPT with a standardized follow-up question following the original question: “Based on the passage’s context, give me your best-educated guess to the above question.”) (“EDUC” strategy)
2. Answer at the same per-section accuracy (simulated by adding the result of the sum of visual item questions in a section multiplied by the per-section accuracy to that section’s raw number of correct answers) (“PSA” strategy)

## RESULTS

### ChatGPT yields good accuracy, achieving median and higher than median performance on the MCAT

Exam items were first encoded as multiple-choice single answers without forced justification (NFJ). This input format is the verbatim question format presented to test-takers. With indeterminate responses excluded/included, ChatGPT accuracy for the MCAT CP, CARS, BB, and PS sections was 58%/50%, 75%/75%, 76%/75%, and 76%/75%, respectively.

Exam items were also encoded as multiple-choice single answers with forced justification (FJ). The input format is the coupling of the verbatim question format along with a request to force ChatGPT to justify its answer selections. With indeterminate responses excluded/included, ChatGPT accuracy for the MCAT CP, CARS, BB, and PS sections was 60%/57%, 74%/74%, 72%/71%, and 80%/79%, respectively (Figure 2).

**Figure 2.**
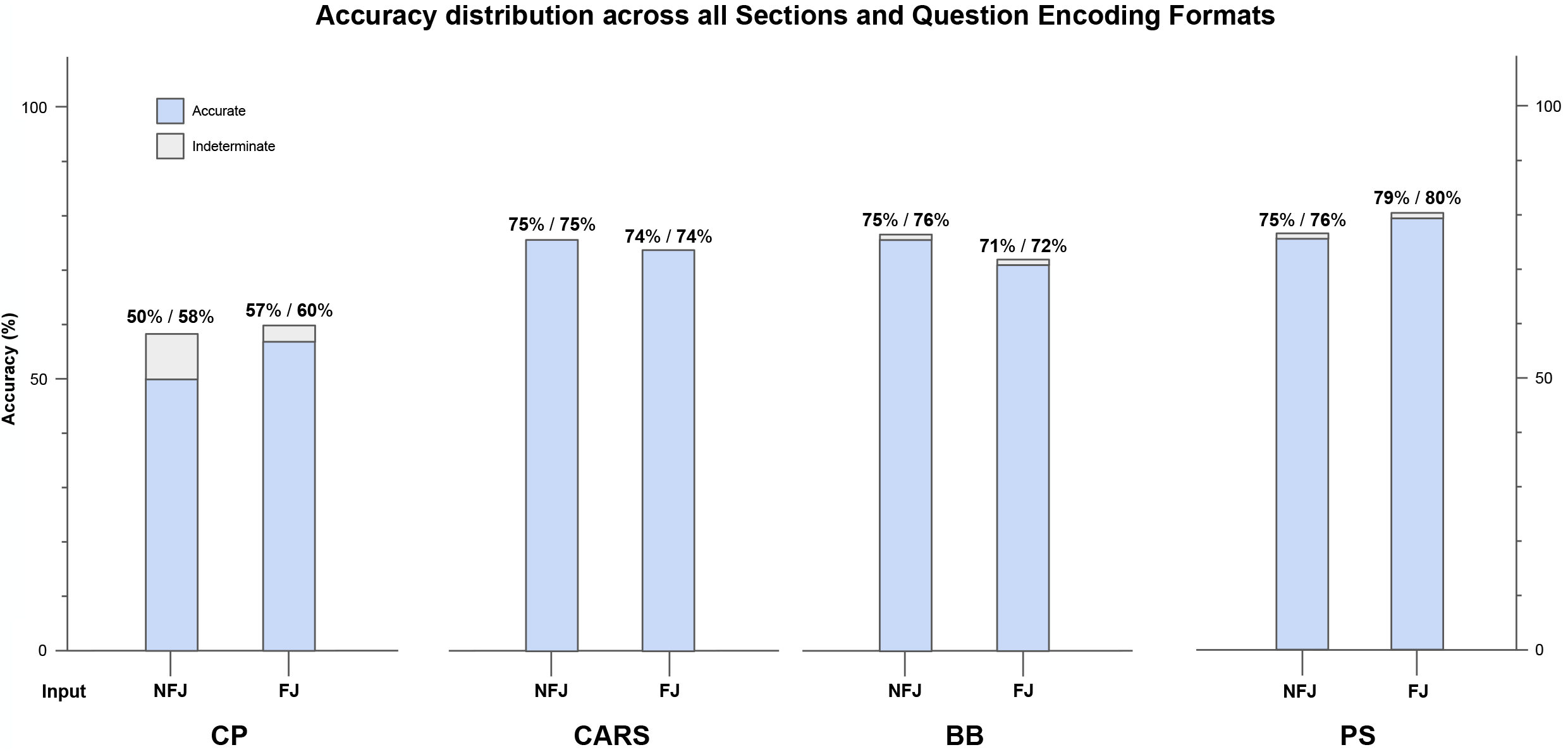
Accuracy distribution for inputs encoded as multiple-choice single answers without (NFJ) or with forced justification (FJ), broken down by section. For CP, CARS, BB, and PS sections, AI outputs were adjudicated to be accurate, inaccurate, or indeterminate based on the scoring system outlined in Table 1.

For the “EDUC” visual item response strategy requiring input into ChatGPT, visual item multiple-choice questions were encoded as NFJ variants. This input is the verbatim question format presented to student test-takers. For CP, CARS, BB, PS, and Total MCAT section scores, ChatGPT’s scaled scores were “EDUC” (125, 126, 125, 126, and 502) and “PSA” (124, 126, 127, 127, and 504), respectively. Under the “EDUC” and “PSA” strategies, ChatGPT performed at or beyond the median total MCAT performance of all 276,779 test-takers from 2019 to 2021 (Figures 3A and 3B).

**Figure 3.**
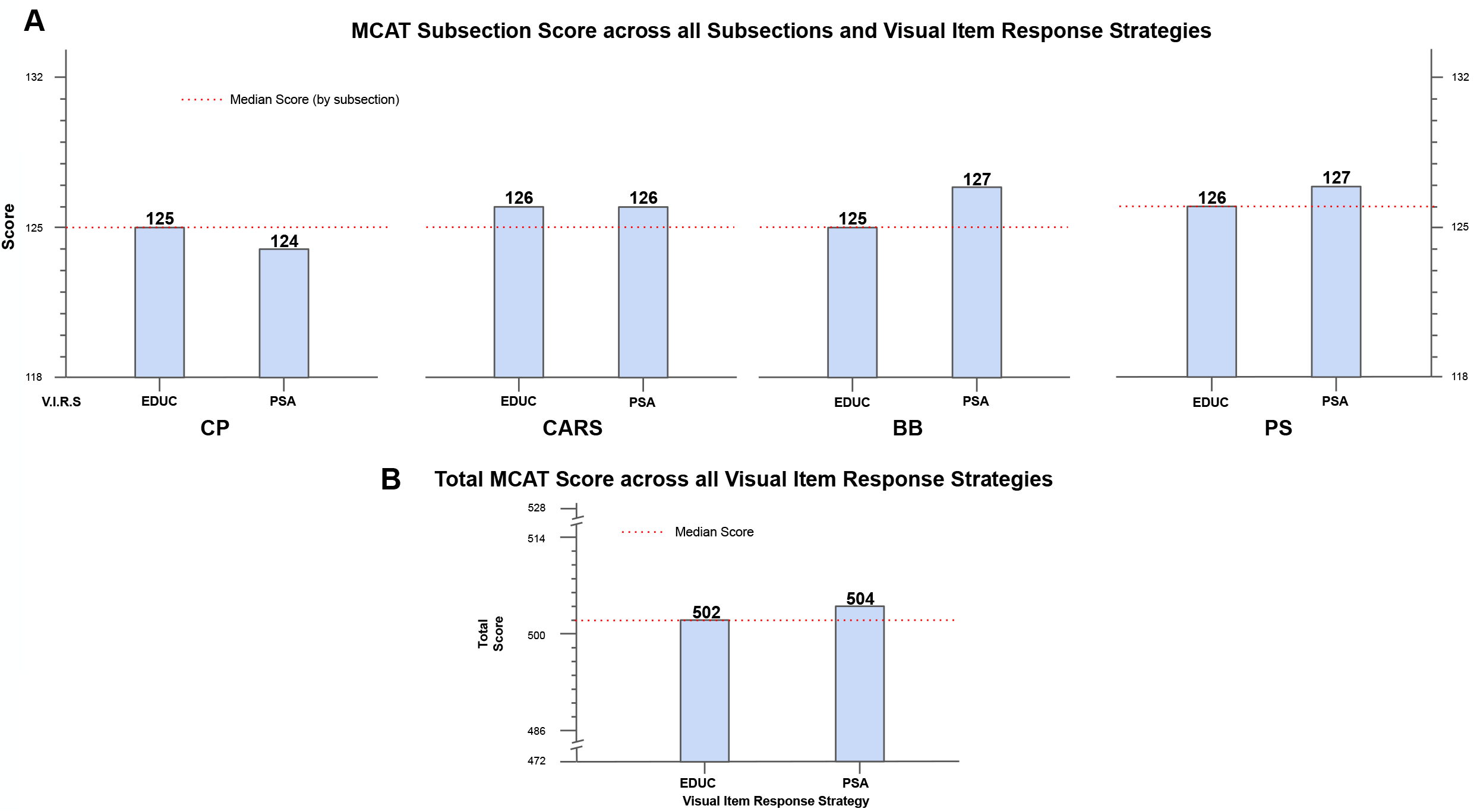
Performance of ChatGPT evaluated as scaled scores. Five visual item response strategies are used to rank ChatGPT’s performance on the MCAT. Multiple choice single answer without forced justification was used as the input encoding format for all 230 questions. Scaled scores were calculated through AAMC system input. The median score (shown by the red dotted line) for both sections and the total score was determined via percentile ranks calculated by the AAMC, based on the rounded median score of the test-taker group (*n* = 276,779) who tested in 2019, 2020, and 2021. **A:** Section scores were calculated across all visual item response strategies. Abbreviation: V.I.R.S = Visual Item Response Strategy. **B:** Total MCAT scores were calculated across all visual item response strategies. “EDUC” refers to the strategy where ChatGPT is prompted to make an educated guess using a standardized prompt. “PSA” refers to the assumption strategy where the sum of visual item questions is multiplied by the per-section accuracy.

### ChatGPT demonstrates high agreement with the official answer key

The agreement with the answer key was adjudicated by an adjudicator via manual inspection of the explanation content based on the scoring system outlined in Table 1. For the NFJ-encoded question variants, ChatGPT outputted explanations for the MCAT CP, CARS, BB, and PS sections with 78%, 81%, 80%, and 75% agreement across all questions, respectively. For the FJ-encoded question variants, ChatGPT outputted explanations for the MCAT CP, CARS, BB, and PS sections with 74%, 92%, 82%, and 88% agreement across all questions, respectively. High agreement was sustained across all exam levels as well as NFJ and FJ question input formats (Figure 4A).

**Figure 4.**
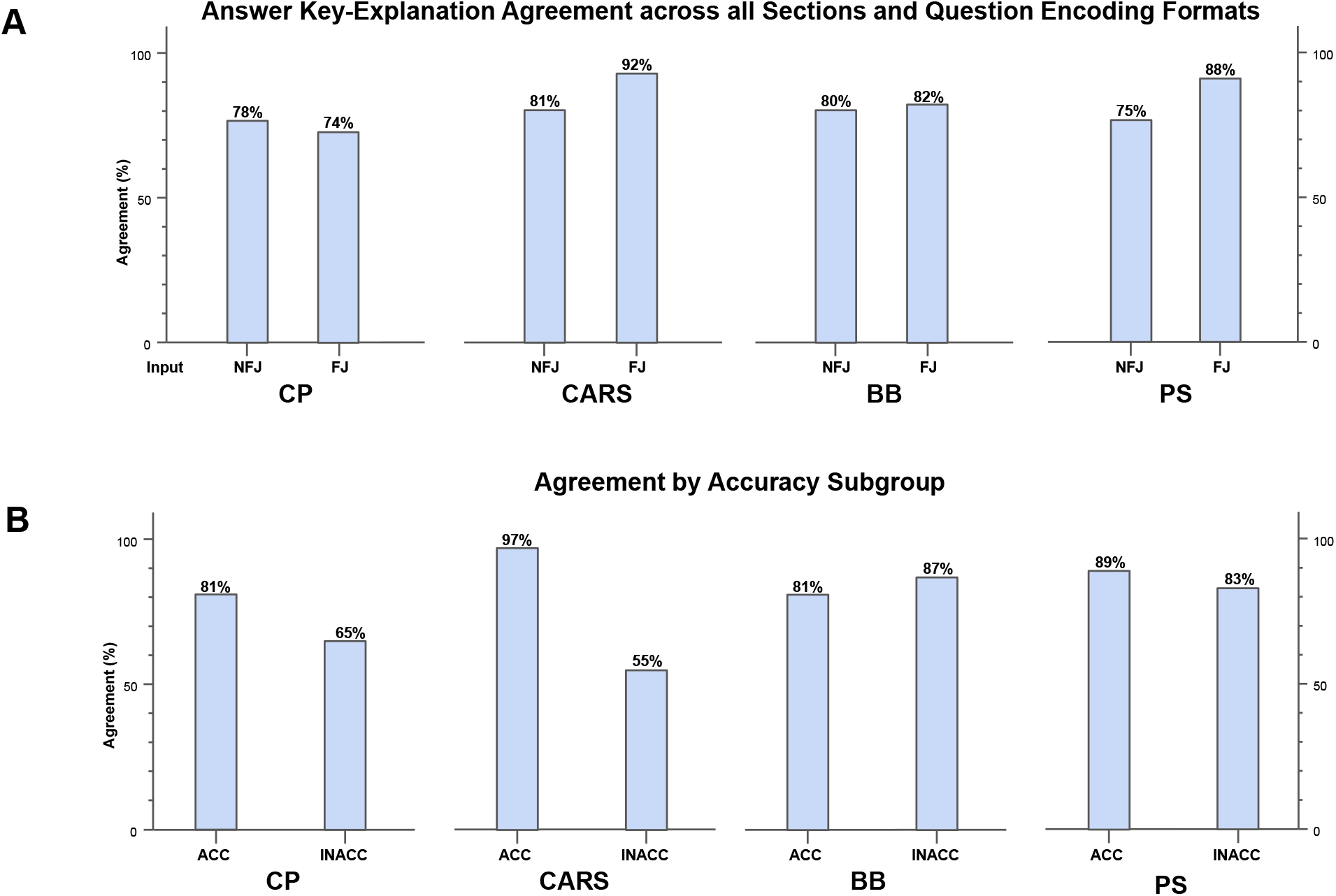
Answer key-explanation agreement of ChatGPT on MCAT. For CP, CARS, BB, and PS sections, AI outputs were adjudicated on their agreement with the answer key based on the scoring system outlined in Table 1. Each instance of answer key-explanation agreement was assigned a value of 1 and disagreement was assigned a value of 0 to enable the calculation of percentile agreement. **A:** Percentile agreement of ChatGPT with the answer key, with questions encoded as multiple-choice single answers without (NFJ) or with forced justification (FJ), broken down by section. **B:** Percentile agreement of ChatGPT with answer key stratified between accurate vs inaccurate outputs, broken down by section. The FJ question encoding format was used.

To analyze if ChatGPT could still generate proper answers whether or not it correctly responded to a question, we analyzed the contingency between accuracy and agreement in FJ-encoded responses, where ChatGPT was forced to justify its answer choice preference. For the MCAT CP, CARS, and PS sections, agreement with the answer key amongst accurate responses was high and greater than amongst inaccurate responses (Figure 4B).

### Insights offered by ChatGPT may assist premedical students

Given ChatGPT’s accuracy and agreement with the established answer key, we next examined its potential to provide insightful answers and thus augment learning. AI-generated explanations were adjudicated by a reviewer. Explanation content was examined for significant insight, defined as the answer choice explanations that met the criteria in Table 1. These analyses were performed on FJ-encoded question variants only.

For the MCAT CP, CARS, BB, and PS sections, ChatGPT produced at least one significant insight in 87%, 85%, 86%, and 91% of all responses, respectively; the prevalence of insight was highly consistent even between content sections that tested disparate competencies (Figure 5).

**Figure 5.**
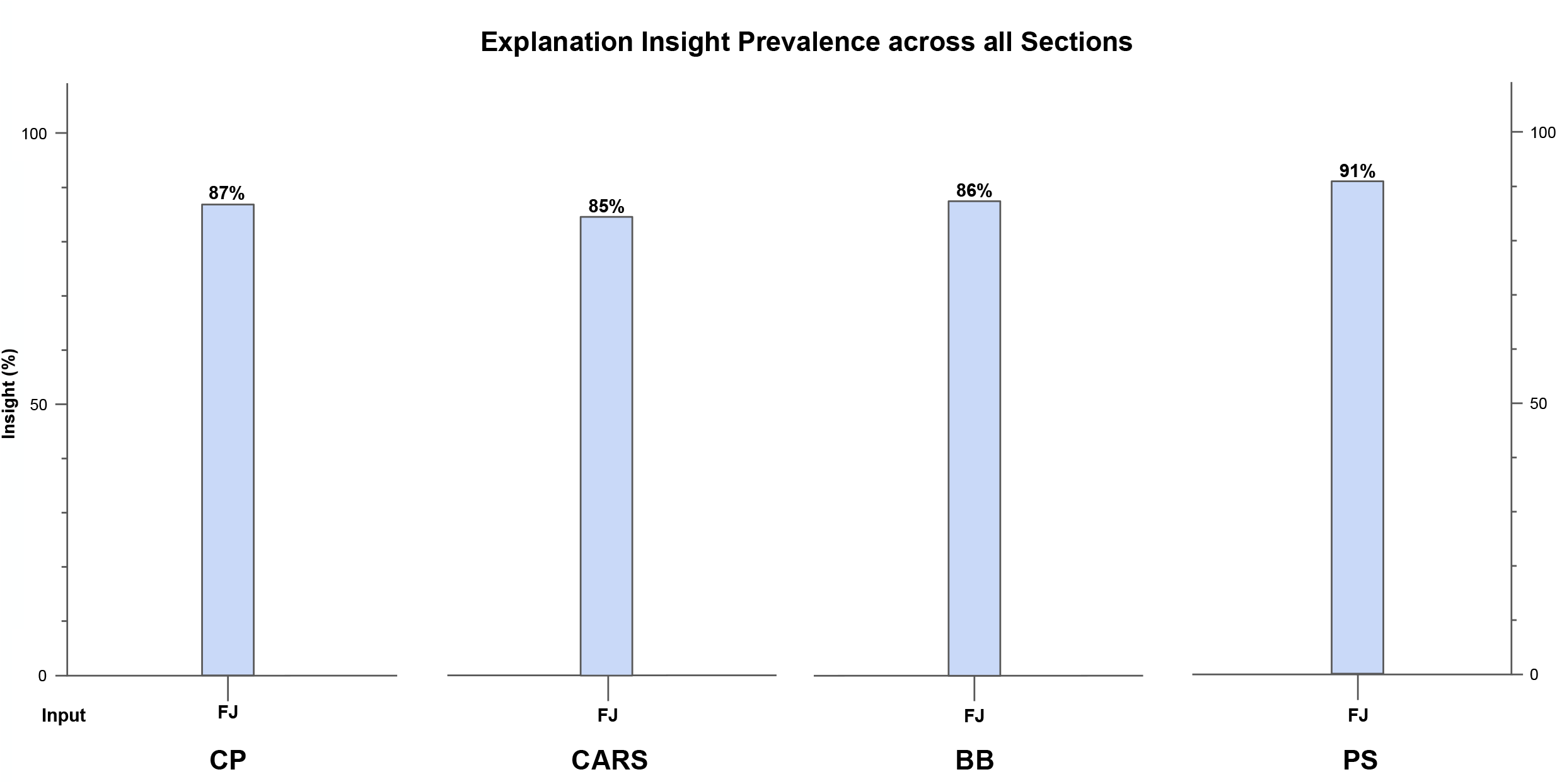
Overall insight prevalence, defined as the proportion of the explanations associated with ChatGPT’s chosen answer with 1 insight, across all exam sections for questions encoded in the FJ format. For CP, CARS, BB, and PS sections, AI outputs were adjudicated on their insight based on the scoring system outlined in Table 1. Each instance of insight was assigned a value of 1 and a lack of insight was assigned a value of 0 to enable the calculation of overall insight prevalence.

Explanations for all possible answer choices were examined to quantify the density of insight contained within AI-generated explanations. High-quality outputs were generally characterized by a DOI of at least 0.5 (i.e. nondefinitional, nonobvious, and valid explanations provided for at least 2 out of 4 wrong answer choices). Across the CP, CARS, BB, and PS sections, we observed that DOI was higher in question items answered accurately versus inaccurately (Figure 6). When answering correctly, these results suggest that a student is likely to gain new insight from the ChatGPT explanations on MCAT multiple-choice questions. Conversely, if answering incorrectly, a student learner is less likely, but still able to gain additional insight.

**Figure 6.**
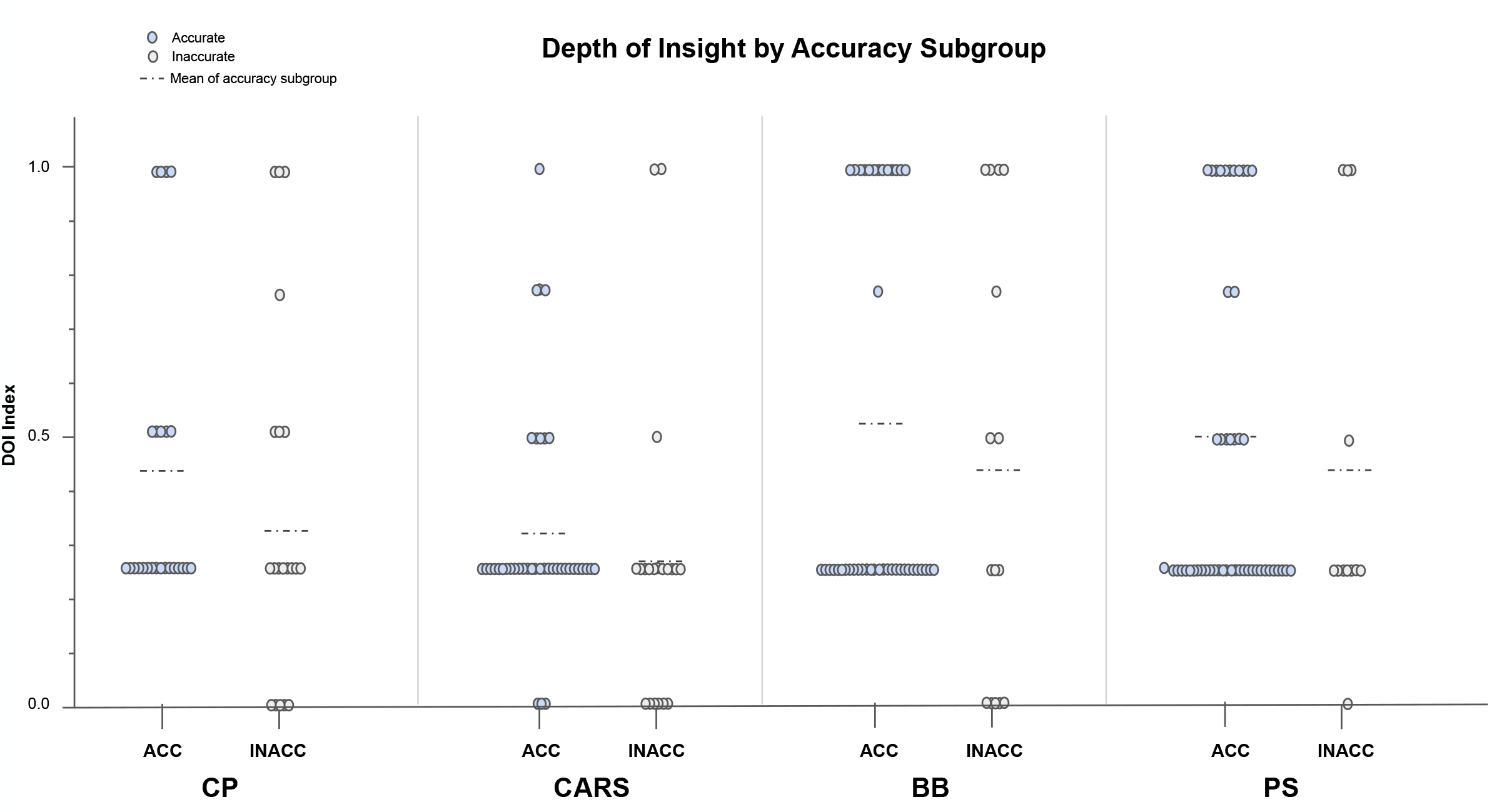
DOI stratified between accurate vs inaccurate outputs, across all exam subsections for questions encoded in FJ format. The DOI metric is calculated by taking the number of ChatGPT’s explanations for each of the possible answer choices that meet the insight criteria (as defined by the scoring system in Table 1) and dividing by 4, the number of possible answer choices on the MCAT. The horizontal dot-dash line indicates the mean of the subgroup. Abbreviation: DOI = Density of Insight.

## DISCUSSION

In this study, we provide new evidence that ChatGPT can perform several tasks related to answering a breadth of standardized questions necessary to be admitted into medical schools. Particularly, to assess its capabilities on pre-medical questions of standardized difficulty, we tested its performance against the MCAT. While previous studies have examined the performance of LLMs on other requisite standardized exams to become a physician (i.e., USMLE STEP 1, STEP 2 CK, and STEP 3), this is the first study examining the performance of an LLM on the MCAT. Our results demonstrate that ChatGPT can achieve a score near or higher than the median scaled score on the MCAT, solidifying its accuracy. Moreover, ChatGPT displays high levels of agreement with the established answer key and insight when prompted for answer justification. Since ChatGPT has demonstrated answer accuracy, agreement with the answer key, and insight on a breadth of questions corresponding to various pre-medical competency areas, our results indicate ChatGPT’s potential utility as a future study tool to augment premedical students’ education in these same competency areas.

Based on these results, we anticipate two primary applications of ChatGPT in premedical education. Firstly, ChatGPT and future advanced iterations of LLMs could provide free access to individualized explanations of MCAT-related materials for all students from all socioeconomic backgrounds, even as early as high school. The issue of underrepresentation of students from URM groups in medicine and even the broader STEM field is systemic, beginning with being less prepared in pre-college and college courses^18^. Less access to and participation in Advanced Placement science courses, lack of support and guidance from family and faculty mentors, and financial challenges can all affect whether a URM student may persist in becoming a physician^19, 20^.

Generating a cohort of medical students and future physicians that reflects the growing racial and ethnic diversity of the country is a crucial goal for medical schools nationwide. Medical school admission officers use MCAT scores and other measures of academic preparation and personal attributes to select the applicants they consider the most likely to succeed in medical school. Due to educational barriers for URMs, this exam poses a barrier to diversity given that Black or Latinx applicants score lower on the MCAT than their Caucasian peers^21^. With further performance improvement, free or even low-cost web-accessible chatbots powered by LLMs, such as ChatGPT, can provide an external didactic resource in pre-college and college education. In our opinion, based on the promising results in this study attesting to ChatGPT’s accuracy and explanation insight, LLMs could potentially increase equitable outcomes for URMs by serving as accessible study tools.

Secondly, ChatGPT could be used to generate additional test questions, either by AAMC test-makers or for targeted preparation by pre-medical students. The pre-medical education system, the process of obtaining a stratifying score through exams, and related test prep services constitute a thriving industry. Although its relevance in selecting students likely to thrive in medical school is still contentious^22^, standardized testing has become a crucial aspect of premedical education. The biggest challenge in creating new exam material is the human capital needed to design practical scenarios that test important concepts, encourage critical thinking, and provide valuable insights regardless of the student’s answer. The demand for practice exam content is continuously growing. Research has shown that increasing the use of full-length MCAT practice exams correlates positively with MCAT performance^23^. Future research may examine if LLMs can relieve some of the laborious human efforts in creating these practice tests by helping with the question-explanation writing process or even producing entire exam passages independently.

Still, despite the promises of this research study, there are still significant limitations. Among the chief limitations is the limited input size of a single MCAT exam, which impacted the scope and depth of our analysis. Moreover, further examination using stratification techniques, such as categorizing the output from ChatGPT based on the subject matter or competency area, could provide valuable information for premedical educators by highlighting any gaps in language processing across heterogenous critical reasoning and content recall tasks. Additionally, questions were consecutively inputted into a single chat window following the prompting of the corresponding MCAT passage. This technique and the predetermined order of the questions could introduce insofar unknown biases by allowing ChatGPT to learn from previous questions, thus creating potential variability in ChatGPT’s response. Although inputting questions consecutively into a single chat window replicates real-world student exam experiences of approaching multiple questions in order associated with a single passage, future studies could consider separating questions by chat window and/or testing different question order permutations within a single chat window to minimize any potential variability in answer-generation caused by ChatGPT’s memory of previously asked questions. A thorough analysis of ChatGPT’s failures, such as obvious language parsing errors in the questions in the CP section, could also offer insight into the source of inaccuracies. Future studies should utilize more sophisticated methods, such as lemmatization, stemming, and other natural language processing techniques, to improve adjudication efficiency and validity. ChatGPT and other similar LLM-powered chatbots must still be tested in real-world learning environments with undergraduate students at varying levels of knowledge and engagement to fully evaluate the effectiveness of LLMs in premedical education.

Nevertheless, our study’s results demonstrate the growing ability of LLMs such as ChatGPT to both accurately approach standardized test questions as well as provide unique insights and explanations into test questions. We suggest that future iterations of these AI software tools may evolve into open-access educational resources that provide equitable, individualized study tools for pre-medical students of all socioeconomic and underrepresented backgrounds, thus paving the way for a more diverse and representative cohort of future physicians.

## Data Availability

All data produced in the present study are available upon reasonable request to the authors.

https://www.mcatofficialprep.org

## ACKNOWLEDGMENTS

The authors thank Joseph Geraghty and Vishal Bommineni for their insightful discussions, guidance, and help in verifying the methodology.

